# Acetylation of Adenine Nucleotide Translocase, Fuel Selection, and Metabolic Flexibility in Human Skeletal Muscle

**DOI:** 10.1101/2022.05.05.22274505

**Authors:** Neusha Barakati, Rocio Zapata Bustos, Dawn K. Coletta, Paul R. Langlais, Lindsay N. Kohler, Moulun Luo, Janet L. Funk, Wayne T. Willis, Lawrence J. Mandarino

**Affiliations:** Division of Endocrinology, Department of Medicine, The University of Arizona, Tucson, Arizona; Center for Disparities in Diabetes, Obesity, and Metabolism, University of Arizona, Health Sciences, Tucson, Arizona; Department of Physiology, The University of Arizona, Tucson, Arizona; Department of Health Promotion Sciences, Mel and Enid Zuckerman College of Public Health, The University of Arizona, Tucson, Arizona; Department of Epidemiology and Biostatistics, Mel and Enid Zuckerman College of Public Health, The University of Arizona, Tucson, Arizona

**Keywords:** Insulin resistance, Metabolic flexibility, mitochondria, Adenine Nucleotide Translocase

## Abstract

**Introduction:** Healthy, resting skeletal muscle primarily oxidizes lipid, but insulin resistant muscle oxidizes carbohydrate and shows metabolic inflexibility during hyperinsulinemia. It is unclear whether fuel selection and metabolic flexibility are dependent on insulin sensitivity in skeletal muscle performing mild exercise.

**Research Design and Methods:** Sedentary volunteers underwent a cycle exercise protocol using stepwise increments in power output (15, 30, and 45 watts) and indirect calorimetry to estimate fuel oxidation in working muscle. Euglycemic clamps, indirect calorimetry and muscle biopsies were used to measure insulin sensitivity and acetylation and content of Adenine Nucleotide Translocase 1 (ANT1), which might be involved in fuel selection via acetylation of lysine 23, which was quantified using mass spectrometry.

**Results:** Mild exercise produced predicted rates of oxygen consumption (11-12 ml O_2_/min), with low and stable blood lactate, allowing use of indirect calorimetry to calculate a respiratory exchange ratio in working muscle (RERm). ANT1 acetylation varied from 0.6 to 21% (10.3 ± 1.2%). Exercising muscle mainly oxidized carbohydrate (45 ± 9, 62 ± 6, and 70 ± 5% of total at 15, 30, and 45watts). Multiple linear regression showed that RERm rose with increasing power output (P < 0.001) and was lower with greater protein content of ANT1 (P < 0.001). Insulin-stimulated glucose disposal, ANT acetylation, and VO_2peak_ were not predictors of RERm.

**Conclusions:** Mildly exercising muscle in sedentary people prefers to oxidize carbohydrate independent of insulin sensitivity but depending on ANT1 protein content. The ability to oxidize lipid may be regulated by higher ANT1 content due to either higher mitochondrial abundance or greater ANT content per mitochondrial mass.

## Introduction

Healthy, resting skeletal muscle prefers to oxidize lipid ^1-5^, but when muscle is exposed to insulin, carbohydrate becomes preferred ^6^. In contrast, insulin resistant resting skeletal muscle prefers to oxidize carbohydrate, and exposure to insulin only modestly raises carbohydrate oxidation ^2-5^. The inability of insulin resistant skeletal muscle to alter the mix of oxidative fuels and its dependence on carbohydrate has been called metabolic inflexibility ^3 7^. In humans, methods to directly study fuel oxidation in skeletal muscle involve arterial and venous catheterization to measure oxygen consumption and carbon dioxide production by a muscle bed and infer the ratio of carbohydrate to lipid oxidation from the Respiratory Quotient (RQ) or calculate rates of fuel oxidation ^3 6 8^. These technical complexities have hindered a more complete characterization of skeletal muscle metabolic inflexibility and its molecular mechanisms. Moreover, although it is evident that metabolic inflexibility characterizes insulin resistant, resting skeletal muscle, less is known regarding whether insulin resistant muscle exhibits metabolic inflexibility during exercise.

Previous studies of metabolic flexibility during exercise have reported only whole-body rates of fuel oxidation and no attempts were made to separate non-muscle from working muscle components of systemic gas exchange. Four studies have assessed whole-body fuel selection during mild to moderate intensity cycling exercise ^9-12^. These studies used whole-body indirect calorimetry and prolonged periods of moderate to intense exercise. Although these studies provide important insight into whole-body fuel selection of obese and insulin resistant subjects, it remains unclear whether working muscle in such individuals oxidizes carbohydrate at different rates than healthy people. Moreover, the mechanisms underlying any such a difference in the fuel selection of active muscle have not been elucidated. Limitations of those studies are that the duration and intensity of exercise is higher than might be considered mild exercise for individuals who are unfit due to long-term physical inactivity and skeletal muscle fuel oxidation is not estimated. Thus, one of the purposes of the present study was to determine whether insulin resistance compromises metabolic flexibility in exercising skeletal muscle is compromised across a range of very mild exercise intensities, which might be more representative of daily routine movement.

Metabolic inflexibility has been well characterized ^3-6 13-20^ but the mechanisms responsible for it are poorly understood ^13^. Mitochondrial ATP production and fuel selection are linked ^21^, and a wide variety of differences in mitochondrial content and function have been described in insulin resistant muscle. The precise nature of differences in mitochondrial function and content in healthy and insulin resistant skeletal muscle are still a matter of investigation. Mitochondria respond to the respiratory signal of [ADP] to ensure demand for ATP is met, and [ADP] is a signal that coordinates ATP production and fuel selection. Rising [ADP] from hydrolysis of ATP results in conditions that favor carbohydrate uptake and oxidation ^22-25^. The mitochondrial inner membrane protein Adenine Nucleotide Translocase (ANT) is the major locus of flux control for respiration and oxidative phosphorylation ^26^. At low rates of ATP turnover, sensitivity of mitochondria to [ADP] mainly depends on the cellular content of mitochondria, ANT abundance, and the binding affinity of ANT for ADP. Moreover, acetylation of ANT1, the predominant isoform of ANT in skeletal muscle ^26^, may play a role in regulation of the sensitivity of mitochondria to [ADP] through acetylation at lysine residue 23 by lowering the overall positive charge of this region of ANT1 and reducing the binding affinity of ADP dramatically ^27 28 29^. Thus, acetylated ANT1 molecules can be viewed as nearly non-functional at [ADP] in resting muscle. Acetylation of lysine 23 of ANT1 therefore would be predicted to lead to a higher requirement for [ADP] needed to meet the energy demands of the cell ^26^, resulting in activation of glycolysis and a greater contribution of carbohydrate to the supply of oxidative fuels ^30 31^. Therefore, the second purpose of this study was to use a novel technique for quantifying ANT1 abundance and acetylation in human skeletal muscle to determine whether these variables predict metabolic flexibility and fuel preference during moderate exercise.

## Methods

### Screening and euglycemic clamps (Figure 1)

**Figure 1.**
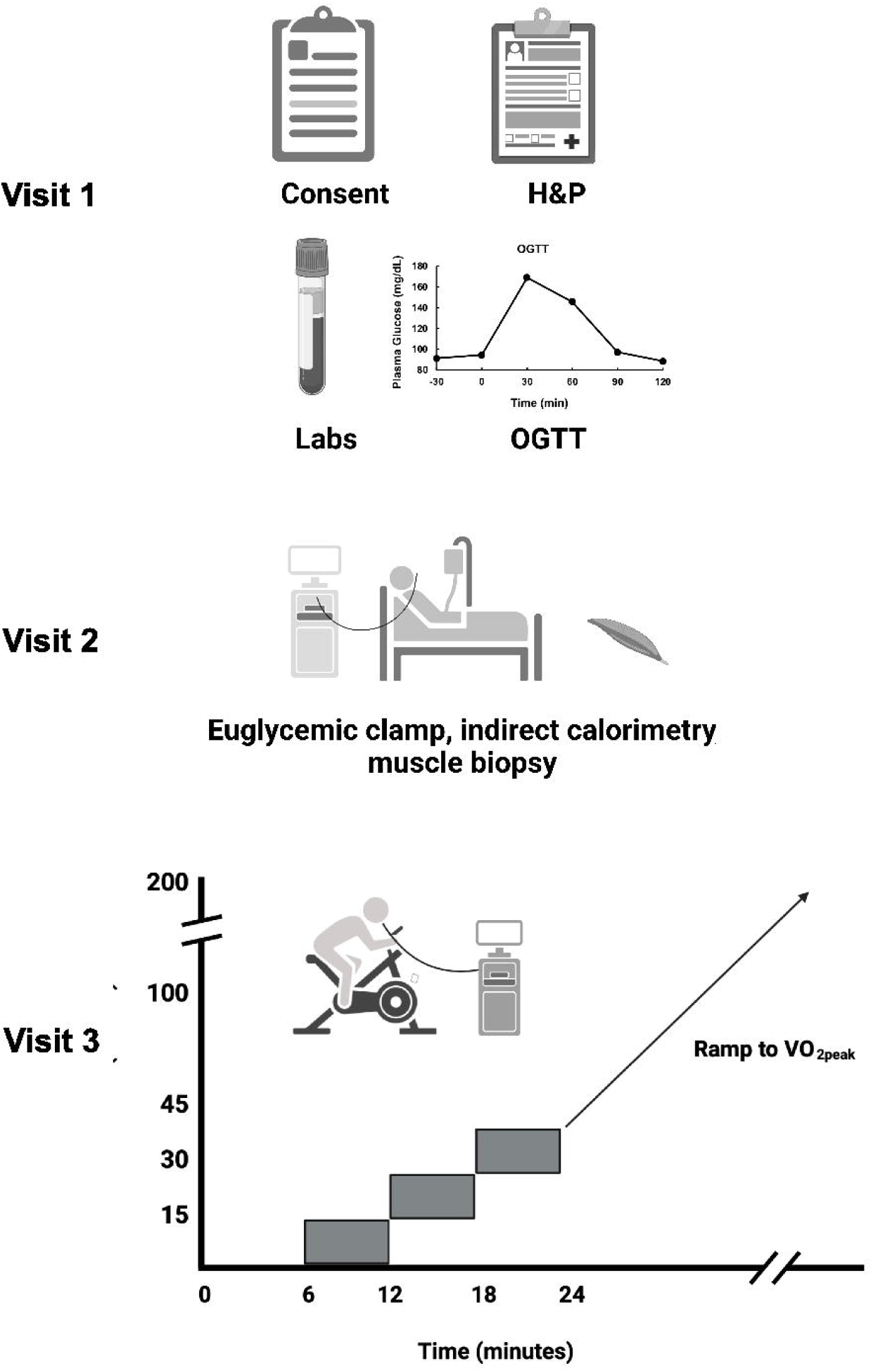
Design of studies. Visit 1 consisted of a screening and consent visit, including history and physical examination, blood drawing for laboratory measurements, and a 75 gram OGTT. Visit 2 consisted of a euglycemic, hyperinsulinemic clamp experiment with muscle biopsies, and Visit 3 consisted of a graded cycle ergometry test with indirect calorimetry and a ramp protocol to determine VO_2peak_.

This study consisted of three visits to the University of Arizona Clinical and Translational Science Research Center, as shown. Studies were approved by the University of Arizona Institutional Review Board, and participants gave written, informed consent. Twenty participants (aged 21-55) received a history and physical examination, screening laboratory measurements, measurement of body composition (bioimpedance), an ECG, and a 75 g oral glucose tolerance test. Participants were not taking any medications that affect glucose metabolism. Participants were instructed to maintain their usual diet and not to engage in exercise 48 hours before any testing.

All volunteers underwent a euglycemic, hyperinsulinemic clamp with indirect calorimetry and percutaneous muscle biopsies, starting at 7-8 am after an overnight fast ^32^. Isotopically labeled glucose (6,6-dideuteroglucose, Cambridge Laboratories) was used to trace glucose metabolism^33^, and steady state conditions were assumed for calculating the rates of glucose metabolism. Participants had a percutaneous needle biopsy of the *vastus lateralis* muscle using under basal conditions one hour before starting an insulin infusion at a rate of 80 mU•m^-2^•min^-1^. One half hour before starting the insulin infusion, indirect calorimetry was used to measure basal rates of oxygen consumption and carbon dioxide production. Plasma glucose was measured every 5-10 minutes and euglycemia was maintained using an infusion of 20% dextrose. Ninety minutes after starting the insulin infusion, indirect calorimetry was again used to measure gas exchange for 30 minutes. At time 120 minutes a second muscle biopsy was performed. Biopsies were frozen in liquid nitrogen for later analysis^32 33^.

### Exercise testing (Figure 1, Visit 3)

On another day, at least one week separated from the glucose clamp, gas exchange measurements were made at rest and during progressive cycle exercise. After catheter placement for blood sampling, subjects rested on the cycle for 6 minutes, then exercised for six minutes periods each at power outputs of 15, 30, and 45 watts. VO_2_ and VCO_2_ values at rest and during the three exercise periods were used to calculate ΔVO_2_ and ΔVCO_2_ during. Delta values were used to calculate a Respiratory Exchange Ratio due to working muscle (RERm = ΔVCO_2_/ΔVO_2_) and rates of substrate oxidation using the equations of Frayn^8^. RERm estimates the oxidative metabolism of muscle performing mild exercise^34^. Blood was collected for measurement of lactate concentrations during this period. This was followed by a ramp protocol to determine VO_2peak_. One subject could not adequately complete the exercise protocol due to anxiety and another had resting blood lactate levels greater than 2 mM, so their exercise data were not used.

### Muscle biopsy processing

Muscle biopsies were stored in liquid nitrogen until they were homogenized for preparation of lysates for immunoblots and measurement of ANT1 acetylation and ANT1 content, as described^32^.

### ANT1 acetylation analysis

ANT is one of the most abundant mitochondrial proteins ^35^. Making it feasible to quantify ANT using lysates of whole muscle. ANT1 is the sole identifiable ANT isoform in human skeletal muscle ^26^. ANT1 acetylation (%) and abundance were estimated using ^13^C and ^15^N-labeled synthetic peptides surrounding the acetylation site (lysine 23) as internal standards ^36^. These synthetic heavy-labeled peptides had the same amino acid sequences as the tryptic peptides surrounding Lys23 that are observed in mass spectrometry experiments ^27^. The abundance of unacetylated ANT1 is referred to as “functional” ANT1 considering the dramatically lower affinity for the acetylated form ^27^.

### Immunoblot analysis

Lysates of muscle biopsies were used for immunoblot analysis of abundance and phosphorylation of serine 232 of pyruvate dehydrogenase (PDH), which lowers the activity of this enzyme ^32^.

### Analytical assays

Enrichment of deuterated glucose was determined by LC-MS ^37^. Insulin was assayed using ELISA (Alpco, Salem, NH). Blood lactate was measured using either Analox (Analox Instruments, Lunenberg, MA) or YSI (YSI, Yellow Springs Instruments) lactate analyzers. Plasma insulin was assayed using ELISA ^33^. Citrate synthase activity was determined as described ^38^.

### Calculations and statistics

Rates of glucose turnover were calculated using steady state equations ^39^. Carbohydrate and lipid oxidation rates were calculated as described ^8^. Statistical comparisons were performed using t-tests or analysis of variance. Pearson’s correlation coefficient was used to assess relationships between two variables. The effects of exercise on blood lactate concentrations and whole-body RER were determined using repeated measures analysis of variance (anova procedure, Stata software, StataCorp, College Station, Texas). The relationships among RERm at submaximal workloads, ANT1 content, ANT1 acetylation, insulin sensitivity (ΔRd), PDH, and VO_2peak_ were analyzed using a mixed linear models approach (xtmixed procedure, Stata).

## Results

### Participant characteristics (Table 1)

Participants (n = 20) had a large range of adiposity (BMI 20.4 – 47.0). Fasting plasma glucose and insulin averaged 89 ± 0.1 mg/dl and 5.8 ± 0.7 µU/ml; HbA1c varied between 4.9 - 6.0%. Recruitment over this range was intentional, to represent a broad range of insulin sensitivity.

**Table 1.** Characteristics of participants.

|  | Mean | SEM |
| --- | --- | --- |
| Age (years) | 31.2 | 2.3 |
| Sex (F/M) | 13/7 |  |
| Weight (Kg) | 76.7 | 3.9 |
| BMI ($\text{Kg/m}^2$) | 26.7 | 1.3 |
| Fat Body Weight (Kg) | 23.8 | 2.3 |
| Lean Body Weight (Kg) | 52.9 | 2.5 |
| Body Fat (%) | 30.5 | 1.7 |
| Hb A1c (%) | 5.3 | 0.1 |
| Glucose (mg/dL) | 88.9 | 2.3 |
| Fasting Serum Insulin (mU/L) | 5.8 | 0.7 |
| HDL (mg/dL) | 51.6 | 2.6 |
| LDL (mg/dL) | 89.5 | 5.1 |
| VLDL (mg/dL) | 19.0 | 1.7 |
| Total Cholesterol (mg/dL) | 160.0 | 5.7 |
| Triglycerides (mg/dL) | 94.5 | 8.7 |
| Resting heart rate (BPM) | 67.3 | 2.5 |
| Systolic blood pressure (mm Hg) | 125.2 | 4.0 |
| Diastolic blood pressure (mm Hg) | 73.8 | 2.4 |
| Waist/Hip Ratio | 0.85 | 0.02 |

Characteristics of volunteers who participated in the study. Data are given as Means ± SEM.

### Glucose metabolism and insulin action (Figure 2)

Responses to 75g oral glucose are shown in Figure 2 A and B. By OGTT criteria ^40^, three participants had type 2 diabetes mellitus, although all three patients had HbA1c between 5.7 and 6.0%, nondiabetic fasting plasma glucose, and were not being treated with diabetes medications. During the glucose clamp, insulin infusion raised serum insulin to 109 ± 7 µU/ml. Basal rates of glucose disposal and appearance were approximately 3.5 mg•Kg-FFM^-1.^min^-1^. Insulin infusion suppressed endogenous glucose production and raised glucose disposal to over 9 mg•Kg-FFM^-1.^min^-1^ (Figure 2C). Rates of insulin-stimulated glucose disposal varied from 3.0 - 17.8 mg•Kg-FFM^-1.^min^-1^. Fat oxidation (Figure 2D) fell during hyperinsulinemia, and carbohydrate oxidation rose from ∼1 to nearly 4 mg•Kg-FFM^-1.^min^-1^ (range 2.2 - 6.7 mg•Kg-FFM^-1.^min^-1^). Basally, 78 ± 3% of energy expenditure was from lipid, falling to 34 ± 4% during hyperinsulinemia. The fraction of fuel oxidation during hyperinsulinemia accounted for by carbohydrate was strongly correlated with insulin stimulated glucose disposal (Figure 2E).

**Figure 2.**
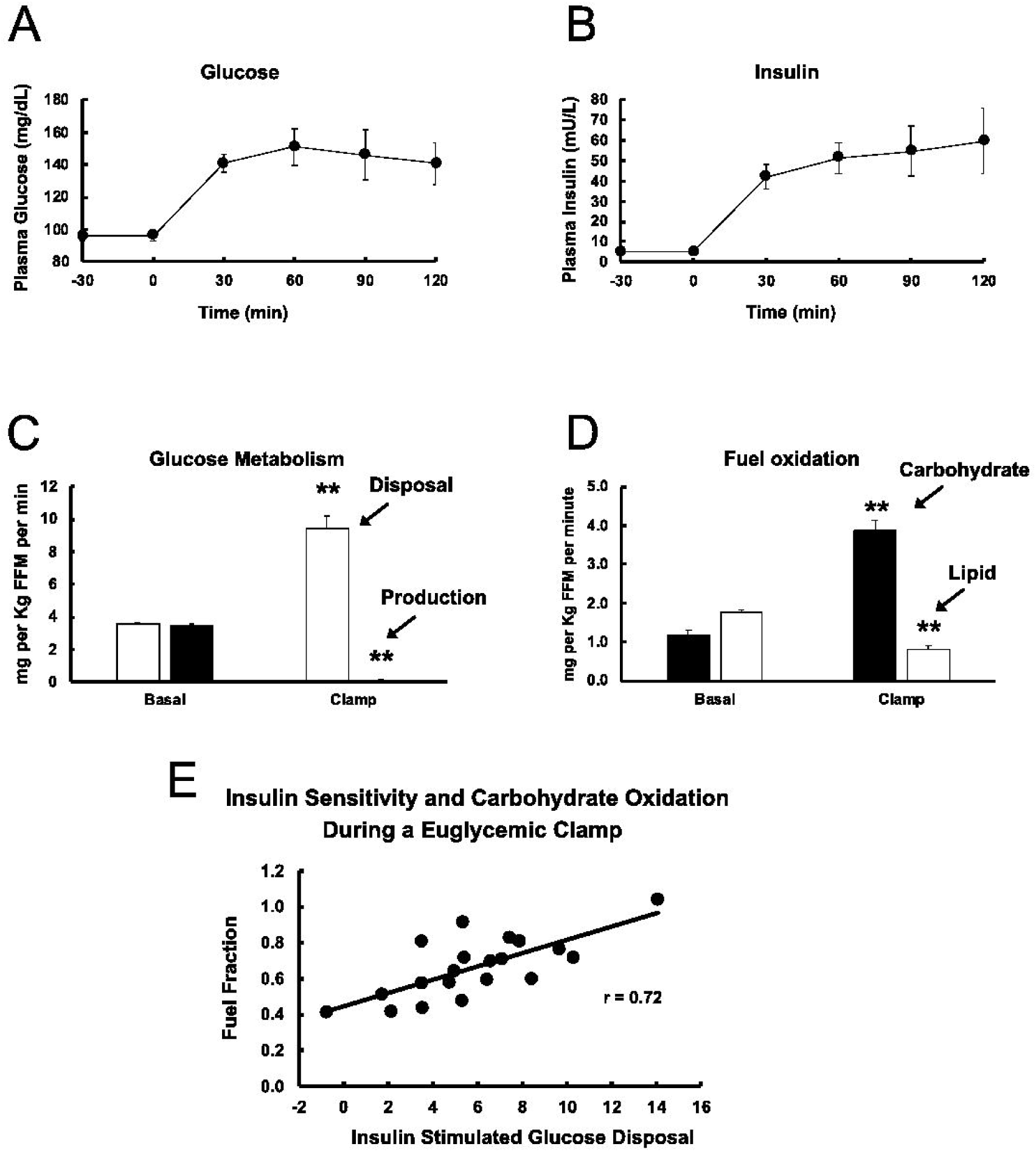
Results of OGTT and glucose clamp studies. Plasma glucose (A) and insulin (B) concentrations following a 75 gram oral glucose load; (C) Rates of glucose disposal (open bars) and endogenous production (closed bars) under basal, postabsorptive conditions and during the glucose clamp; (D) Rates of carbohydrate (closed bars) and lipid (open bars) oxidation determined by indirect calorimetry basally and during the glucose clamp; (E) correlation between insulin stimulated glucose disposal and the fraction of fuel oxidation accounted for by carbohydrate. Data are given as Means ± SEM. **P < 0.01 vs. basal.

### Validation of exercise protocol and effect of exercise on fuel selection in working muscle

Volunteers underwent an exercise study with whole-body gas exchange at rest and during mild steady state exercise of 15, 30, and 45 watts, followed by a ramp protocol to VO_2peak_. Changes in VO_2_ and VCO_2_, between rest and exercise are mainly attributable to fuel oxidation in active muscle^34^, allowing calculation of the respiratory exchange ratio due to muscle, or RERm ^34 41^. The validity of this approach to assess fuel oxidation in working muscle using systemic indirect calorimetry was tested by examining whole-body RER, blood lactate, the linearity and slope of the relationship between VO_2_ and power output, and mechanical efficiency of cycling. Whole-body RER fell from a resting value of 0.81 to about 0.78 at 15 watts and then rose to 0.86 at 45 watts of exercise. Blood lactate rose modestly from about 0.85 at rest to about 1.40 mM at 45 watts of exercise (Figure 3A). Lactate concentrations were stable during the final 3 minutes of each 6-minute period of exercise at 15, 30, or 45 watts. The relationship between oxygen consumption and power output was linear, with an O_2_ cost of approximately 11.8 ml O_2_/min/watt power (Figure 3B). The relationship between power output and whole-body energy expenditure rate also was linear, with a slope indicating a cycling efficiency of about 24%. Absolute power outputs represented relative efforts of about 10, 20, and 30 percent of maximum power achieved at VO_2_peak, or 29 ± 1, 36 ± 2, and 45 ± 2 percent of whole-body oxygen consumption at VO_2peak_ at 15, 30, and 45 watts, emphasizing the mild nature of this exercise. RERm (RER due to working muscle) is shown in Figure 3C at each power output. Repeated measures analysis of variance shows a rise in RERm with increasing power output (F = 11.33, df = 2, P = 0.002), indicating a rise in carbohydrate oxidation as a percent of fuel use. Corresponding rates of carbohydrate and lipid oxidation and energy expenditure from the two fuels during exercise are given in Table 2. Following steady state exercise, a ramped exercise protocol was used to estimate VO_2peak,_ which averaged 34.7 ml•Kg-FFM^-1^ (range 21.7 - 54.5 ml•Kg-FFM^-1^) and was a significant predictor of insulin stimulated glucose disposal (r = 0.55, P < 0.01, Figure 3D).

**Figure 3.**
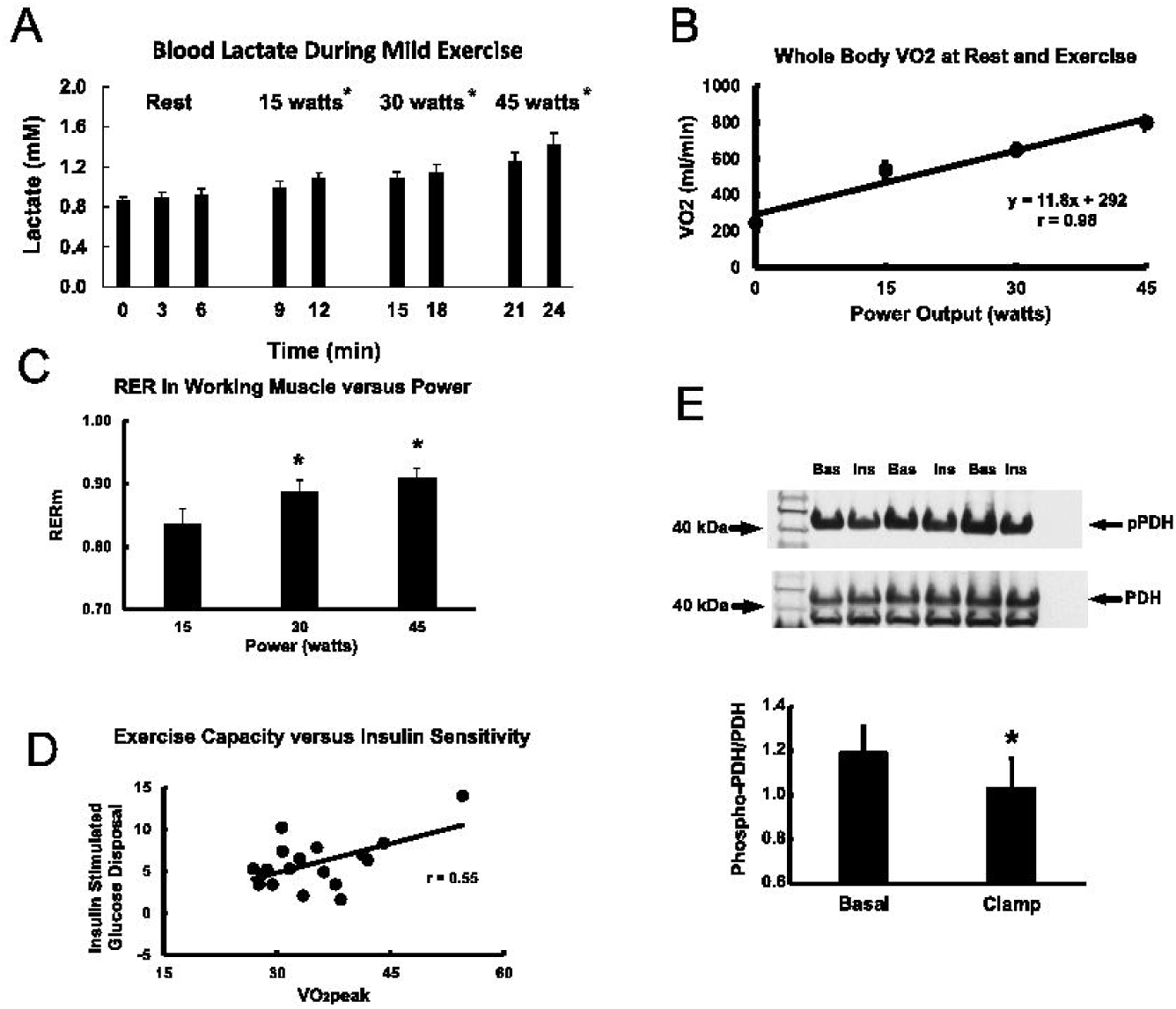
Validation of whole-body indirect calorimetry for estimating Respiratory Exchange Ratio in mildly exercising skeletal muscle. (A) Blood lactate concentrations (mM) at rest and at each three-minute period of exercise at 15, 30, and 45 watts (6 minutes total exercise at each power output); B) Linear relationship between power output and whole-body VO_2_; (C) RER due to working muscle (RERm) at three power outputs; (D) Positive linear relationship between VO_2peak_ and insulin sensitivity; (E) Representative immunoblots of phosphorylated PDH and total PDH protein (top panel), and quantification of the effect of insulin (clamp) to lower phosphorylation of PDH. Data are given as Means ± SEM. *P < 0.05 vs. rest or insulin.

**Table 2.** Fuel oxidation rates and caloric expenditure during mild exercise.

|  | Power Output in watts (% maximum) |  |  |
| --- | --- | --- | --- |
| | 15 ($9.8 \pm 0.6$) | 30 ($19.5 \pm 1.2$) | 45 ($29.3 \pm 1.9$) |
| Fuel |  |  |  |
| Carbohydrate, $\text{mg} \cdot \text{min}^{-1}$ | $150 \pm 36$ | $319 \pm 40^{**}$ | $493 \pm 43^{**}$ |
| Carbohydrate, $\text{kcal} \cdot \text{min}^{-1}$ | $600 \pm 144$ | $1914 \pm 160^{**}$ | $1972 \pm 172^{**}$ |
| Lipid, $\text{mg} \cdot \text{min}^{-1}$ | $79 \pm 15$ | $77 \pm 14$ | $83 \pm 15$ |
| Lipid, $\text{kcal} \cdot \text{min}^{-1}$ | $711 \pm 135$ | $693 \pm 126$ | $747 \pm 135$ |

Rates of fuel oxidation are given as Mean ± SEM in units of mg/min. Caloric values are provided as $\text{kcal} \cdot \text{min}^{-1}$ . Rates were calculated from $\Delta VO_2$ and $\Delta CO_2$ during exercise according to Frayn<sup>8</sup>. $^{**}P < 0.01$ vs 15 watts.

*ANT1 acetylation*. Acetylation of lysine 23 of ANT1 in muscle lysates averaged 10.5 ± 1.1% (range of 0.6 to 21.2%). ANT1 content was 7.07 ± 0.42 nmoles/gram muscle wet weight (range 2.66-11.2 nmoles/gram wet weight). Functional ANT1, the unacetylated fraction of ANT1, was 6.29 ± 0.37 nmoles/gram wet weight (range 2.33 - 9.94 nmoles/gram).

### Phosphorylation of pyruvate dehydrogenase (PDH) is regulated by insulin

PDH activity is regulated in part by phosphorylation at serine 232 of the E1 subunit. A representative immunoblot of PDH phosphorylated at serine 232 and total PDH protein is given in Figure 3E, upper panel. Infusion of insulin lowered phosphorylation of PDH relative to total PDH protein from 1.21 ± 0.12 to 1.07 ± 0.12 (Figure 3E, lower panel, P < 0.019, paired t-test).

### Relationships among fuel selection during moderate exercise and insulin sensitivity, VO_2peak_, ANT1 acetylation and ANT1 content, and PDH phosphorylation and protein

Mixed model regression analysis for repeated measures was used to assess whether insulin-stimulated glucose disposal, ANT1 acetylation, or functional ANT1 content were independent predictors of RERm during moderate cycling exercise. The overall model was highly significant (P<0.0001). Power output (β= 0.00271 ± 0.0006, P < 0.001), functional ANT1 content (β= -0.0339 ± 0.012, P = 0.004), and basal PDH protein content (β= 7.0 × 10^−5^ ± 3.4 × 10^−5^, P = 0.038) were significant independent predictors of fuel use in working skeletal muscle. The negative β for functional ANT1 content indicates that greater functional ANT1 content predicts lower RERm (greater fat oxidation.) When the rate of lipid oxidation at 45 watts (Table 2) was compared with functional ANT1 content, it was found that the two were directly related (r = 0.62, P < 0.01). Because ANT1 content could be a marker for mitochondrial abundance, we compared ANT1 content and citrate synthase activity in a subset of 12 muscle biopsies that had sufficient material remaining after other assays. ANT1 content per gram wet weight muscle was correlated with citrate synthase activity per gram muscle (r = 0.68, P < 0.05).

## Discussion

The concept of metabolic flexibility ^7 13 20 42^ grew from characterization of the “Randle Cycle” ^43-46^. Healthy, resting skeletal muscle prefers to oxidize lipid and responds to insulin by switching fuel preference to carbohydrate ^1 6^. In contrast, insulin resistant muscle under basal, resting conditions prefers to oxidize carbohydrate ^2 3 5^. Metabolic inflexibility refers to reduced ability of resting skeletal muscle of insulin resistant obese and type 2 diabetic patients to appropriately select oxidative fuels. With dietary caloric oversupply and reduced caloric expenditure, lipid accumulates ectopically in muscle and likely contributes to insulin resistance ^13^. As originally applied in the context of insulin resistant muscle, metabolic flexibility specifically referred to a high basal rate of carbohydrate oxidation with little if any change brought about by raising plasma insulin concentrations during a glucose clamp experiments. Whether exercising, insulin resistant skeletal muscle appropriately selects oxidative fuel is less well-characterized.

Exercise or muscle contraction leads to an increase in the rate of glucose taken up by working muscle ^47^. Different mechanisms for increasing glucose uptake operate during exercise versus insulin stimulation, raising the question of whether fuel selection during mild exercise is related to insulin sensitivity. To answer this question, we took advantage of the fact that changes in systemic gas exchange during mild exercise are mainly due to changes in fuel oxidation in working muscle ^34^. To implement this approach, volunteers engaged in three consecutive periods of cycle exercise at power outputs of 15, 30, and 45 watts. These power outputs on a cycle ergometer translated to a range of 10-30% of maximum power output, or 20-45% of whole-body VO_2peak_. During these periods of mild exercise, increases in blood lactate were small and lactate concentrations were stable during the portion of each 6-minute period where RERm was calculated. During this experiment, the cycling efficiency (about 24%) and oxygen consumption per watt (increment of about 11-12 ml O_2_.min) were within expected values ^47^, so this approach appears to be appropriate for estimating fuel oxidation in mildly exercising muscle.

A mixed model linear analysis using power output, insulin-stimulated glucose disposal rates, VO_2peak_, ANT acetylation, functional ANT content, PDH phosphorylation and PDH protein as predictors of RER in working muscle (RERm) was used to determine the best predictors of fuel choice during mild exercise. The results of this analysis showed that power output was a strong predictor of RERm, indicating that, across a very mild range of exercise intensity, the percent contribution of carbohydrate to energy production rose, while the relative contribution of lipid fell (P < 0.001). However, insulin-stimulated glucose disposal was not a significant predictor of RERm during exercise. Therefore, unlike exposure of insulin resistant muscle to insulin, mild exercise produces changes in fuel oxidation that are independent of insulin sensitivity. There are several earlier studies that addressed the question of metabolic flexibility in exercising muscle. The main finding of the present study, that fuel choice during mild exercise is unaffected by insulin resistance confirms the findings of Colberg and colleagues ^10^. The present studies and those of Colberg^10^ emphasize that, in comparison to more physically active individuals^34^, carbohydrate is the preferred oxidative substrate during mild exercise. Two other studies found lipid oxidation to be higher in subjects who were likely to be insulin resistant^9 11 12^. However, both of those studies used a substantially higher exercise intensity (50% of maximum) and duration (60-90 minutes) than either the current study or that of Colberg ^10^, which might explain the differences. However, it would seem likely that higher intensity exercise would result in a greater contribution of carbohydrate to the fuel mix. Moreover, whole-body RER was measured during those studies, compared to RER in working muscle. Braun and colleagues used a milder exercise protocol (45% max) but for a longer duration and also found higher lipid oxidation during exercise^9^. The duration of exercise and the fact that whole-body RER was used without correction for metabolism in non-working muscle and other tissues may partially explain the differences between those results and the present findings. This gives weight to the argument that ANT content or mitochondrial abundance is a key factor in determining the proportions of oxidative fuels used by mild to moderately exercising skeletal muscle.

Although insulin sensitivity across a broad range was not a predictor of fuel choice in the present studies, that does not imply that the relative contributions of carbohydrate and lipid to oxidative metabolism in muscle was at all “normal”. The present findings, as well as those described above in similar subjects stand in contrast to the findings of Willis *et al*.^34^ in healthy, active individual sand those of Romijn *et al*. ^48^who examined the effects of exercise intensity in endurance-trained women and showed that at mild to moderate exercise intensities, highly trained people have much lower whole-body RER than the participants in the current study or others cited above. These results demonstrate another important finding, namely, that older, sedentary people with very low aerobic capacity (VO_2peak_), even in the absence of disease, depend primarily upon carbohydrate as the main energy source during exercise that is equivalent to mild walking, compared with younger, more active individuals^34^. This implies that when mild walking exercise is recommended to sedentary people, their perception of effort and the difficulty with which they sustain walking exercise is such that such exercise recommendations may serve to discourage people from physical activity.

Although metabolic flexibility during mild exercise appears to be independent of insulin sensitivity or aerobic capacity, there still was substantial variability among individuals in their capacity to shift the mix of carbohydrate and lipid oxidized. To examine this on a more mechanistic level, we used a novel mass spectrometric method^36^ to quantify ANT1 acetylation and abundance in muscle biopsies. Factors that raise the apparent Km ADP for oxidative phosphorylation will raise the [ADP] needed to meet energy demands in resting or moderately exercising skeletal muscle. [AMP] and [Pi] also would rise under such circumstances along with the increase in [ADP]. The outcome of this would be stimulation of glycolysis and PDH activity, favoring carbohydrate oxidation. Modeling data show that the sensitivity of mitochondria to the ADP signal for respiration is determined in part by the level of acetylation of ANT1 at lysine at lysine 23^27^, one of three positive charges responsible for binding ADP to ANT^29^. Acetylation removes one positive charge, resulting in a predicted 30 to 50-fold fall in the binding affinity for ADP. In this study, ANT acetylation at lysine 23 averaged approximately 10-11% (range 5 - 20%). Computational modeling data suggests that acetylation at lysine 23 would need to reach about 25% to have significant effects on the apparent Km ADP for respiration^49 50^. Consistent with this, the present findings show that ANT acetylation was not a significant predictor of changes in RERm during moderate exercise. However, the content of ANT1 in muscle that was not acetylated, that is, ANT1 that would be functional at physiological [ADP], was significantly associated with RERm. These results show that a higher content of functional ANT1 was significantly associated with lower carbohydrate and higher lipid oxidation. Therefore, ANT1 abundance appears be related to fuel selection during mild exercise in the absence of an effect of ANT acetylation. This could be the case if ANT1 abundance were a surrogate for mitochondrial content or if there were higher ANT abundance per unit of mitochondria, or both. To address the former, we assayed citrate synthase activity in a subset of muscle biopsies and found a strong positive correlation between citrate synthase activity, a marker of mitochondrial content, and ANT1 abundance. Higher mitochondrial content results in greater sensitivity of mitochondria to [ADP]^51-53^. Because 80-90% of flux control over respiration or oxidative phosphorylation resides at ANT^26^ it also is likely that a rise in ANT content per mitochondrial mass might have similar effects. Taken together, the results of this study suggest that higher mitochondrial content, and perhaps ANT1 abundance, predict that muscle will prefer to oxidize lipid during mild exercise.

PDH phosphorylation and total protein were assessed by immunoblot analysis basally and after insulin infusion to test the prediction that higher sensitivity to [ADP] would be associated with inactivation of PDH via higher phosphorylation by PDH kinase. In this study, only protein content of PDH was significantly, albeit modestly, associated with RERm. This suggests that only a minor portion of control of fuel selection resides at PDH. This leads to the speculation, that regulation of glycolysis may be more important than regulation of PDH during mild exercise.

Taken together, the results of this study show that metabolic flexibility is not lost in insulin resistant muscle performing mild work, like walking. The difference between the effects of exercise and the impact of insulin on skeletal muscle fuel selection may be due to differences in fuel availability during these two conditions, the divergent molecular mechanisms by which muscle contraction and insulin lead to increased glucose transport and uptake, and the need to increase energy expenditure to meet energy demand during muscle contraction. Mechanistically, acetylation of ANT1 at lysine 23 does not appear to be high enough to significantly affect the sensitivity of mitochondria to [ADP] and influence fuel selection. However, higher ANT1 abundance in skeletal muscle is a good predictor of a greater ability to rely on lipid oxidation during exercise, reflecting either higher ANT1 abundance, greater mitochondrial content, or greater ANT1 content per mitochondrial mass.

## Data Availability

All data produced in the present study are available upon reasonable request to the authors

## Acknowledgements

The authors declare no conflicts of interest. The authors thank Jean Finlayson for expert technical assistance, Judy Krentzel and Alma Leon for nursing assistance, and the volunteers. This study was supported by NIH grant R01DK047936 (LJM) and the University of Arizona Health Sciences.

